# Similar Rates of AKI during the First Two Waves of COVID-19 in Northern Italy: a single-center study

**DOI:** 10.1101/2021.06.13.21258862

**Authors:** Gaetano Alfano, Silvia Giovanella, Francesco Fontana, Jovana Milic, Giulia Ligabue, Francesco Giaroni, Camilla Ferri, Martina Montali, Andrea Melluso, Morisi Niccolò, Giacomo Mori, Riccardo Magistroni, Erica Franceschini, Andrea Bedini, Giacomo Cuomo, Margherita DiGaetano, Marianna Meschiari, Cristina Mussini, Gianni Cappelli, Giovanni Guaraldi

## Abstract

**Introduction:** Two waves of COVID-19 cases have overwhelmed most European countries during 2020. It is unclear if the incidence of acute kidney injury (AKI) has changed during the COVID-19 outbreaks. This study aims to evaluate the differences in incidence, risk factors and outcome of AKI in patients with SARS-CoV-2 infection during the first and second wave of COVID-19.

**Method:** We reviewed the health medical records of 792 consecutive patients with COVID-19 hospitalized at the University Hospital of Modena, Italy, from February 25 to December 14, 2020.

**Results:** AKI was diagnosed in 122 (15.4%) patients. Incidence of AKI remained steady rate during wave-1 (15.9%) and wave-2 (14.7%) (P=0.89). AKI patients were older (P=<0.001) and had a more severe respiratory impairment (PO2/FO2) (P=≤0.001) than their non-AKI counterparts. AKI led to a longer hospital stay (P=0.001), complicated with a higher rate of ICU admission. COVID-19-related AKI was associate with 59.7% of deaths during wave-1 and 70.6% during wave-2. At the end of the period of observation, 24% (wave-1) and 46.7% (wave-2) of survivors were discharged with a not fully recovered kidney function. Risk factors for AKI in patients with COVID-19 were diuretics (HR=5.3; 95%CI, 1.2-23.3; P=0.025) and cardiovascular disease (HR, 2.23; 95%CI, 1.05-5.1; P=0.036).

**Conclusion:** The incidence of AKI (about 15%) remained unchanged during 2020, regardless of the trend of COVID-19. AKI occurred in patients with severe COVID-19 symptoms and was associated with a higher incidence of deaths than non-AKI patients. The risk factors of COVID-19-related AKI were diuretic therapy and cardiovascular disease.

## Introduction

Since the first outbreak in China, COVID-19 has overwhelmed Europe with two “waves” in 2020, leading to a considerable morbidity and mortality burden. [1]

It is currently unclear if the numerous implementations of health care interventions have changed the risk profile of COVID-19 associated complications in hospitalized patients. Acute kidney injury (AKI) is one of the most fearsome complications of SARS-CoV-2 infection. It is associated with a significantly higher rate of intensive care unit (ICU) admission, mechanical ventilation and death compared to COVID-19 patients without AKI.[2–4] The rate of AKI in patients with COVID-19 is extremely variable across countries and may differently affect patients with similar disease severity.[5] In a large cohort of vulnerable patients in the USA, the incidence of AKI approached 60%.[6] In Europe, few studies have reported information on AKI in the general population. The available data estimate a rate of AKI in non-severely ill patients ranging between 4.5%-22% [7–10]whereas in China the overall incidence was about 5.4%. [11]

On this background, Palevsky raised the following thoughtful query “Why has there been this high degree of variability in the reported rates of kidney involvement?”[12] To date, there are no data to provide an evidence-based answer to this question because multiple factors are in play, including patients selection, different care management and spectrum of disease severity. With the aim of understanding the epidemiological changes in the rate of AKI during 2020, we evaluated a cohort of COVID-19 patients hospitalized in a University Hospital in Northern Italy.

## Material and methods

We enrolled all hospitalized patients with COVID-19 who were admitted at the University Hospital of Modena between February 25 and December 14, 2020. The diagnosis of COVID-19 was performed through reverse transcriptase-polymerase chain reaction (RT-PCR). We excluded patients aged <18 years (*n*=2), patients on dialysis (*n*=5), and patients who were had missing serum creatinine on admission (*n*=19). All patients included in the first and second waves were hospitalized for the development of severe symptoms of COVID-19 symptoms. All the enrolled patients were discharged or died at the end of the follow-up.

The demographic characteristics, clinical data, and outcomes were extracted from electronic medical records. AKI was defined according to the Kidney Disease: Improving Global Outcomes (KDIGO) criteria[13]. Baseline serum creatinine (sCr) was defined as sCr at admission. Recovery from AKI was defined as a return to a baseline sCr that did not meet AKI stage 1 criteria[14]. Continuous variables were compared using a sample t-test and Mann-Whitney U test. ANOVA test was used to evaluate differences between three groups of patients.

Categorical data were compared using the χ2 test or Fisher’s exact test. ANOVA test was used to evaluate difference between three groups

Kaplan Meier curves were used to compare survival in AKI and non-AKI patients.

Univariate and multivariate Cox regression analysis determined risk factors for AKI. Variables with a p-value□<□0.01 were included in the multivariate model.

The study was approved by the regional ethical committee of Emilia Romagna (n. 0013376/20)

## Results

Study population included 792 COVID-19 patients subdivided into wave-1 (n=389), interwave (n=57) and wave-2 group (n=346). Mean age of wave-1, interwave and wave-2 was 65.2, 63.7 and 66.3 years old, respectively. At presentation, wave-1 and wave-2 group showed similar clinical characteristics, principally characterized by a mild-moderate respiratory distress (PO_2_/FO_2_ about 250) requiring oxygen supplement in about 70% of cases. Conversely, patients admitted during the interwave period manifested a less severe pulmonary involvement (P=<0.001), ICU admission (P=0.001) and showed a trend toward a better outcome (P=0.055). (Table 1). During the study period 122 cases of AKI were diagnosed. A similar incidence of AKI occured during wave-1 (15.9%), interwave (15.8%) and wave-2 group (14.7%) during hospitalization (P=0.89) (Fig.1, Supplementary Material).

**Figure 1S:**
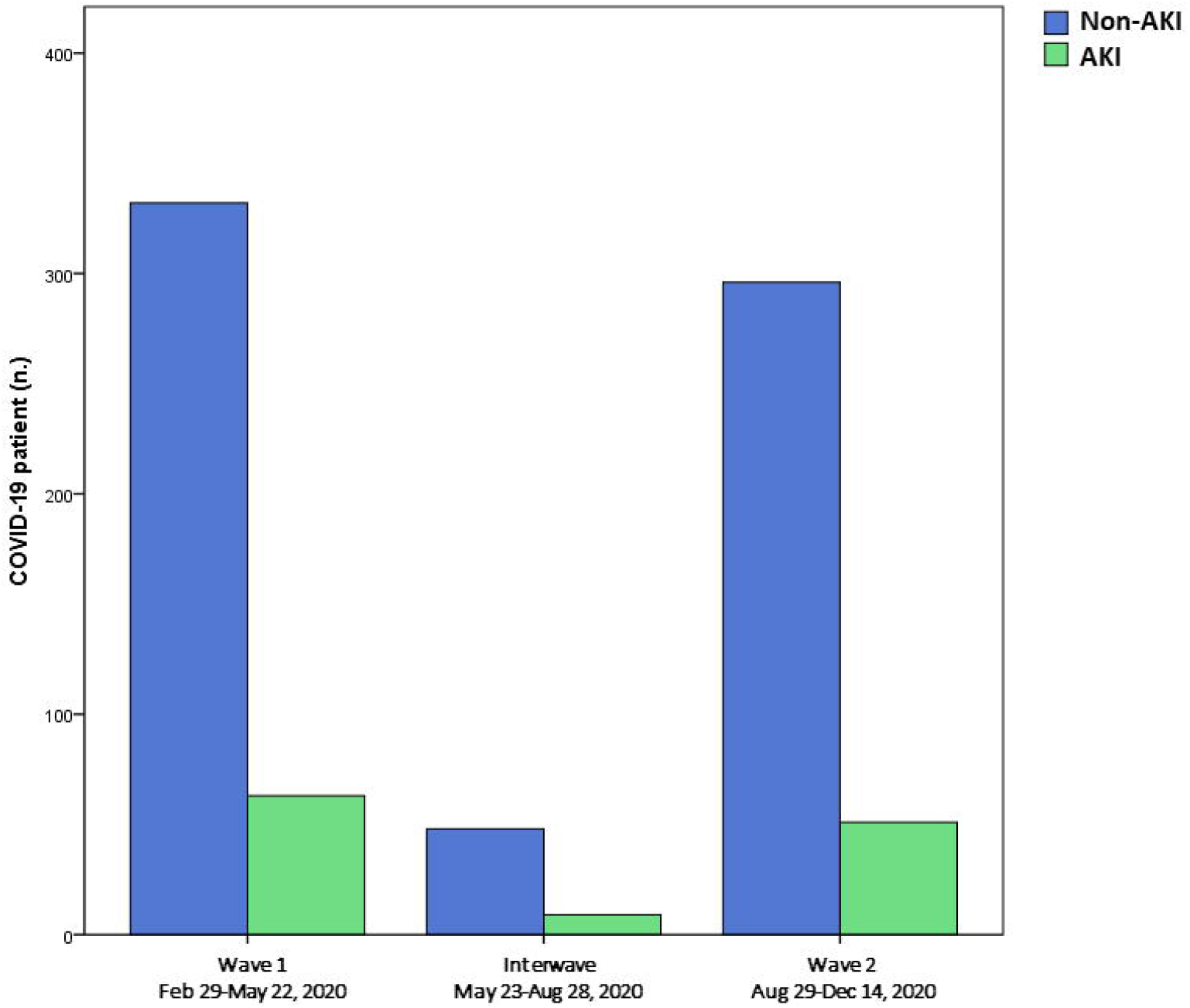
Percentage of AKI and non-AKI patients during wave 1-2.

**Figure 2S:**
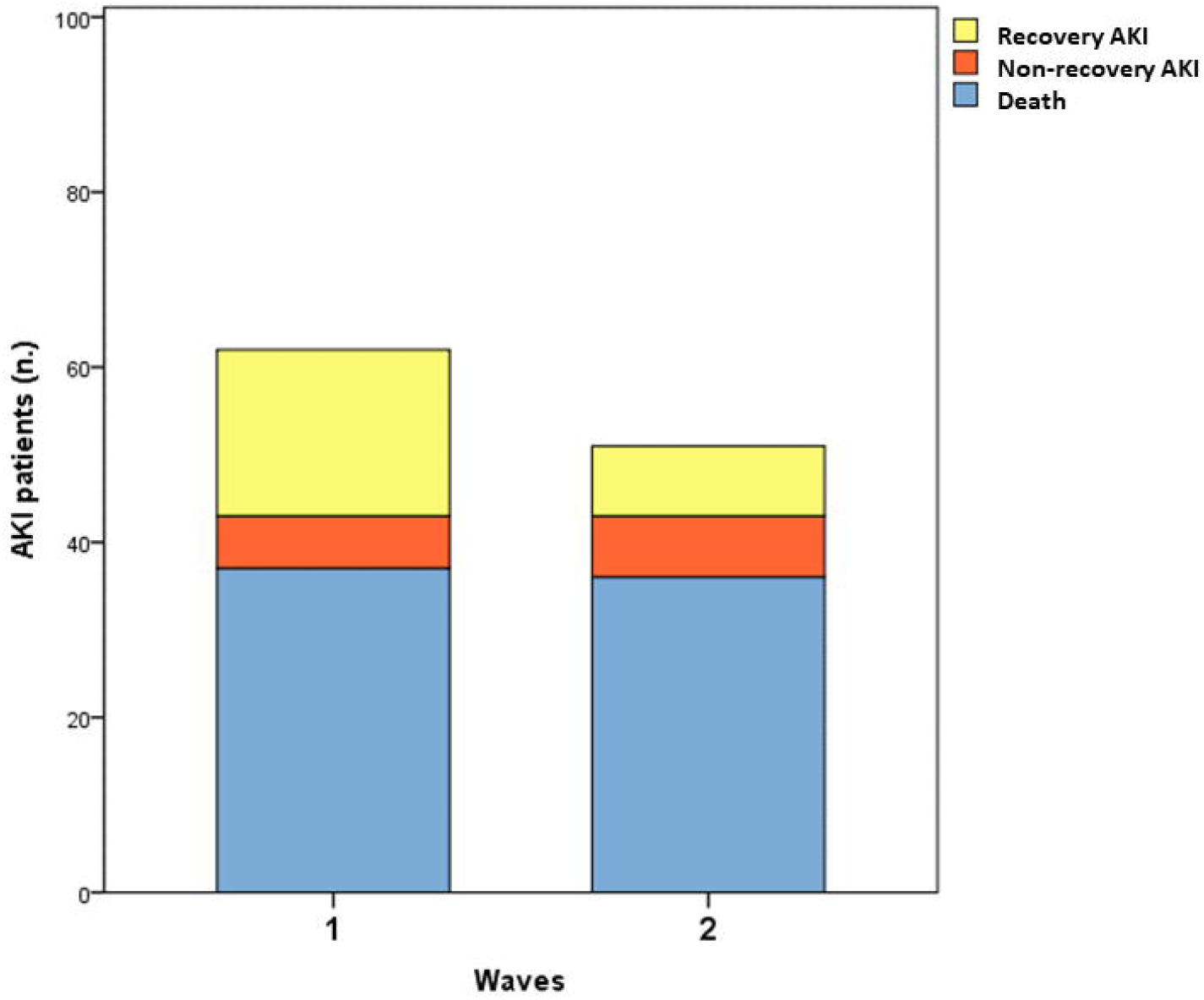
Percentage of survivors (with or without recovery of kidney function) and non-survivors in patients who experienced AKI.

**Figure 3S:**
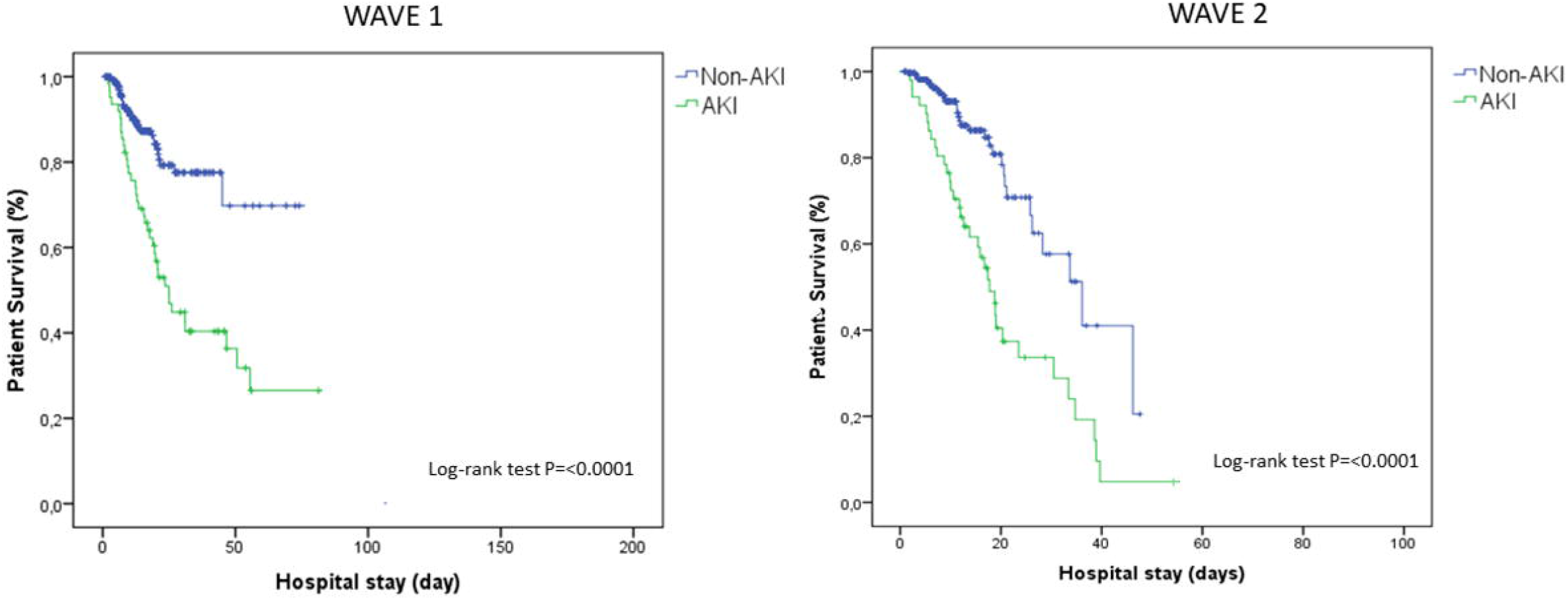
Kaplan-Meier curves of AKI and non-AKI patients stratified according to wave-1 and 2.

**Table 1.** Demographics and clinical manifestation of COVID 19 patients.

| Variable | WAVE-1 Group<br>(n=389) | INTERWAVE Group<br>(n=57) | WAVE-2 Group<br>(n=346) | *p-value* |
| --- | --- | --- | --- | --- |
| Age | 65.2±14.7* | 63.7±22.2 | 66.3±16.1* | **0.024** |
| Males (%) | 262 (67.3) * | 28 (49.1) *<sup>#</sup> | 221 (64)<sup>#</sup> | **0.028** |
| White blood cells (cell/mm<sup>3</sup>) | 8587±7170 | 7022.2±3332 | 7256±4219 | 0.17 |
| Hemoglobin (gr/dl) | 12.6±1.8 | 12.0±2<sup>#</sup> | 13.1±1.6<sup>#</sup> | **0.001** |
| Platelets (10<sup>3</sup>/mm<sup>3</sup>) | 221.9±103.9 | 226.5±102.9 | 220.4±90.0 | 0.924 |
| CRP (mg/dl) | 4.5±6.5<sup>#</sup> | 4.9±7.7<sup>#</sup> | 6.4±7.8<sup>#</sup> | **0.009** |
| LDH (U/L) | 710±1289* | 456.4±202.7* | 589.4±278.5 | **0.021** |
| sCr at admission (mg/dl) | 1.0±0.6 | 0.9±0.6 | 1.0±0.7 | 0.665 |
| MAP | 88.8±12.2* | 86.6±15.8<sup>#</sup> | 95.5±14.2*<sup>#</sup> | **<0.001** |
| PO<sub>2</sub>/FO<sub>2</sub> | 242.5±103.9* | 342±91.5*<sup>#</sup> | 246.7±102.9<sup>#</sup> | **<0.001** |
| SOFA score | 2.1±2 | 1.4±1.6 | 2.1±1.9 | 0.59 |
| Comorbidities<sup>§</sup> (%) | 82.2 | 83.3 | 82.4 | 0.996 |
| COPD (%) | 20/119 (16.8) | 4/10 (36.3)<sup>#</sup> | 8/91 (8.8)<sup>#</sup> | **0.017** |
| Diabetes (%) | 45/131 (34.8) * | 5/8 (62.5) *<sup>#</sup> | 25/96 (26.0) | **0.07** |
| Hypertension (%) | 113/159 (71.1) * | 9/10 (90) *<sup>#</sup> | 60/107 (56.1) | **0.011** |
| CVD (%) | 36/121 (29.8) | 3/7 (42.8) <sup>#</sup> | 11/94 (11.7) <sup>#</sup> | **0.003** |
| CKD (%) | 19/119 (16) | 3/8 (33.3) | 13/94 (13.8) | 0.212 |
| BMI>30 (%) | 36/119 (30.8) | 5/10 (50) | 31/98 (31.6) | 0.436 |
| O<sub>2</sub> therapy (%) | 280 (71.8) * | 21 (36.8) *<sup>#</sup> | 236 (68) <sup>#</sup> | **<0.001** |
| HFNO (%) | 14 (5) | 2 (9.1) | 85 (35) | **<0.001** |
| NIV (%) | 104 (36.2) | 2 (9.1) | 66 (27.4) | **0.007** |
| Mechanical ventilation (%) | 71 (19.9) | 1 (1.7) | 19 (5.4) | **<0.001** |
| Steroid (%) | 77 (21.6) <sup>#</sup> | 11 (19.3) | 199 (57.3) <sup>#</sup> | **<0.001** |
| Immunotherapy (%) | 180 (50.6) | 12 (21.1) | 134 (40) | **<0.001** |
| AKI (%) | 62 (15.9) | 9 (15.8) | 51 (14.7) | 0.89 |
| Hospitalization (days) | 17.4±16.5* | 14.9±10.9 | 11.5±9.2* | **<0.001** |
| **ICU admission (%)** | 94 (26.4) * | 2 (3.5) *<sup>#</sup> | 57 (17.1) <sup>#</sup> | **<0.001** |
| **Death (%)** | 77 (19.7) * | 4 (7) *<sup>#</sup> | 70 (20.2) <sup>#</sup> | 0.055 |

CRP, C-reactive protein; LDH, lactate dehydrogenase; MAP, mean arterial pressure; sCr, serum creatinine; SOFA, Sequential Organ Failure Assessment; COPD, chronic obstructive pulmonary disease; CVD, cardiovascular disease; CKD, chronic kidney disease; BMI, body mass index; HFNO, high-flow nasal oxygen; NIV, noninvasive ventilation; AKI, acute kidney injury; ; ICU, intensive care unit. .
§50% data missing

To better detail the clinical characteristics of AKI population, these patients were compared with respective non-AKI patients (Table 2). The limited number of AKI events in the interwave period (n=9) excluded this group from further analysis.

**Table 2.**
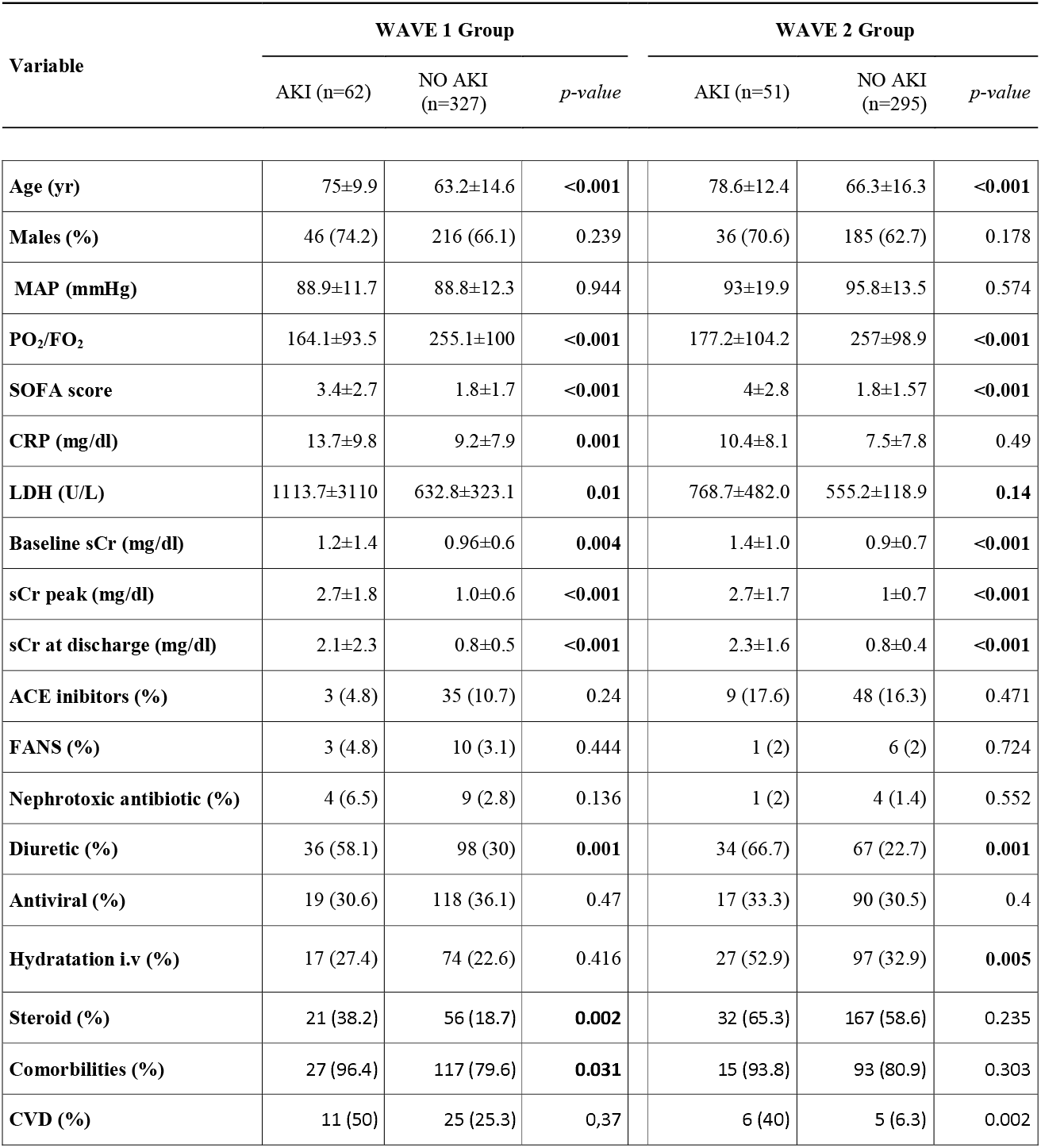
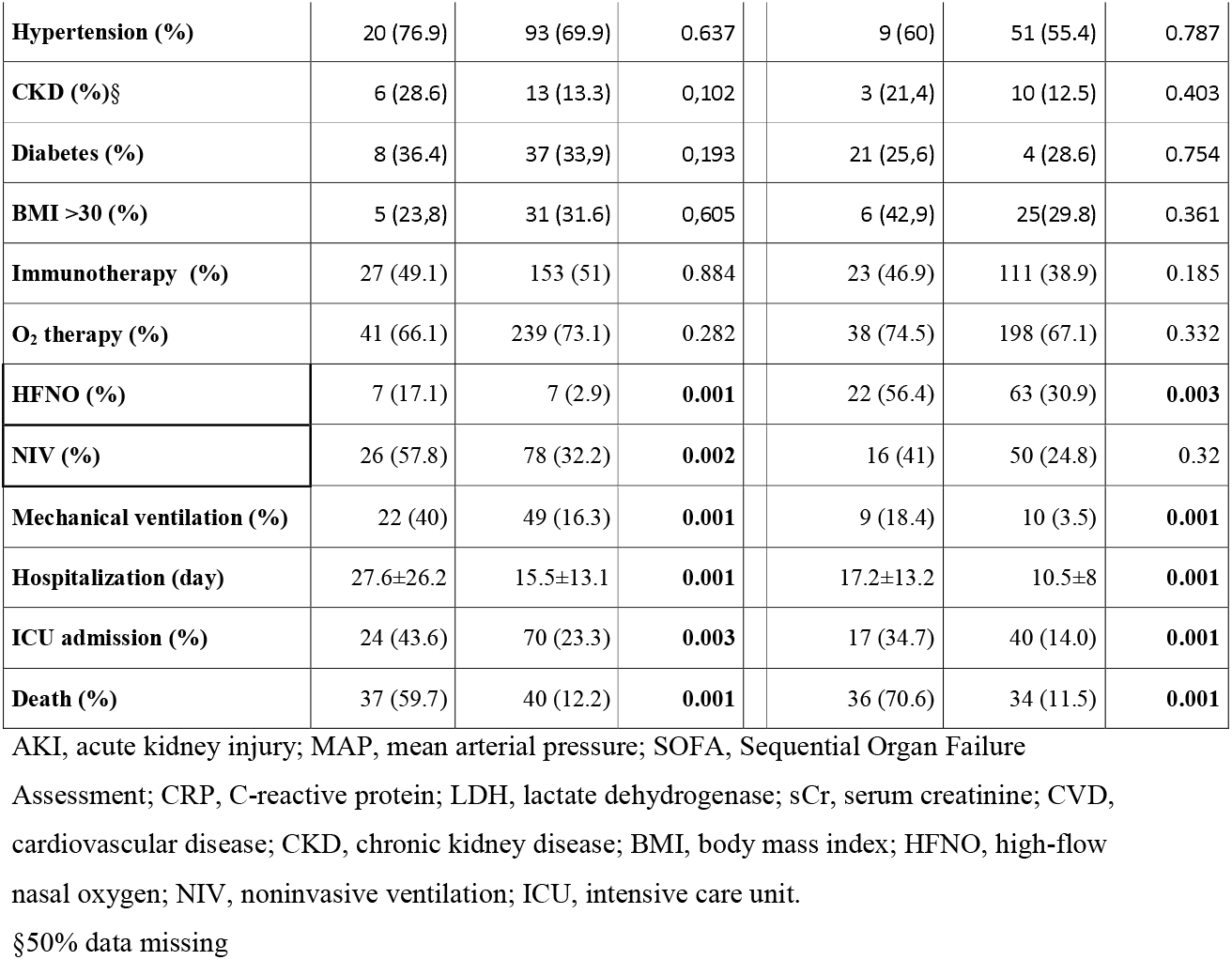
Comparison between AKI and non-AKI patients admitted during wave 1 and 2.

| Variable | WAVE 1 Group |  |  | WAVE 2 Group |  |  |
| --- | --- | --- | --- | --- | --- | --- |
|  | AKI (n=62) | NO AKI (n=327) | *p-value* | AKI (n=51) | NO AKI (n=295) | *p-value* |
| Age (yr) | 75±9.9 | 63.2±14.6 | **<0.001** | 78.6±12.4 | 66.3±16.3 | **<0.001** |
| Males (%) | 46 (74.2) | 216 (66.1) | 0.239 | 36 (70.6) | 185 (62.7) | 0.178 |
| MAP (mmHg) | 88.9±11.7 | 88.8±12.3 | 0.944 | 93±19.9 | 95.8±13.5 | 0.574 |
| PO<sub>2</sub>/FO<sub>2</sub> | 164.1±93.5 | 255.1±100 | **<0.001** | 177.2±104.2 | 257±98.9 | **<0.001** |
| SOFA score | 3.4±2.7 | 1.8±1.7 | **<0.001** | 4±2.8 | 1.8±1.57 | **<0.001** |
| CRP (mg/dl) | 13.7±9.8 | 9.2±7.9 | **0.001** | 10.4±8.1 | 7.5±7.8 | 0.49 |
| LDH (U/L) | 1113.7±3110 | 632.8±323.1 | **0.01** | 768.7±482.0 | 555.2±118.9 | **0.14** |
| Baseline sCr (mg/dl) | 1.2±1.4 | 0.96±0.6 | **0.004** | 1.4±1.0 | 0.9±0.7 | **<0.001** |
| sCr peak (mg/dl) | 2.7±1.8 | 1.0±0.6 | **<0.001** | 2.7±1.7 | 1±0.7 | **<0.001** |
| sCr at discharge (mg/dl) | 2.1±2.3 | 0.8±0.5 | **<0.001** | 2.3±1.6 | 0.8±0.4 | **<0.001** |
| ACE inhibitors (%) | 3 (4.8) | 35 (10.7) | 0.24 | 9 (17.6) | 48 (16.3) | 0.471 |
| FANS (%) | 3 (4.8) | 10 (3.1) | 0.444 | 1 (2) | 6 (2) | 0.724 |
| Nephrotoxic antibiotic (%) | 4 (6.5) | 9 (2.8) | 0.136 | 1 (2) | 4 (1.4) | 0.552 |
| Diuretic (%) | 36 (58.1) | 98 (30) | **0.001** | 34 (66.7) | 67 (22.7) | **0.001** |
| Antiviral (%) | 19 (30.6) | 118 (36.1) | 0.47 | 17 (33.3) | 90 (30.5) | 0.4 |
| Hydration i.v (%) | 17 (27.4) | 74 (22.6) | 0.416 | 27 (52.9) | 97 (32.9) | **0.005** |
| Steroid (%) | 21 (38.2) | 56 (18.7) | **0.002** | 32 (65.3) | 167 (58.6) | 0.235 |
| Comorbidities (%) | 27 (96.4) | 117 (79.6) | **0.031** | 15 (93.8) | 93 (80.9) | 0.303 |
| CVD (%) | 11 (50) | 25 (25.3) | 0.37 | 6 (40) | 5 (6.3) | 0.002 |

|  |  |  |  |  |  |  |  |
| --- | --- | --- | --- | --- | --- | --- | --- |
| **Hypertension (%)** | 20 (76.9) | 93 (69.9) | 0.637 |  | 9 (60) | 51 (55.4) | 0.787 |
| **CKD (%)§** | 6 (28.6) | 13 (13.3) | 0,102 |  | 3 (21,4) | 10 (12.5) | 0.403 |
| **Diabetes (%)** | 8 (36.4) | 37 (33,9) | 0,193 |  | 21 (25,6) | 4 (28.6) | 0.754 |
| **BMI >30 (%)** | 5 (23,8) | 31 (31.6) | 0,605 |  | 6 (42,9) | 25(29.8) | 0.361 |
| **Immunotherapy (%)** | 27 (49.1) | 153 (51) | 0.884 |  | 23 (46.9) | 111 (38.9) | 0.185 |
| **O<sub>2</sub> therapy (%)** | 41 (66.1) | 239 (73.1) | 0.282 |  | 38 (74.5) | 198 (67.1) | 0.332 |
| **HFNO (%)** | 7 (17.1) | 7 (2.9) | **0.001** |  | 22 (56.4) | 63 (30.9) | **0.003** |
| **NIV (%)** | 26 (57.8) | 78 (32.2) | **0.002** |  | 16 (41) | 50 (24.8) | 0.32 |
| **Mechanical ventilation (%)** | 22 (40) | 49 (16.3) | **0.001** |  | 9 (18.4) | 10 (3.5) | **0.001** |
| **Hospitalization (day)** | 27.6±26.2 | 15.5±13.1 | **0.001** |  | 17.2±13.2 | 10.5±8 | **0.001** |
| **ICU admission (%)** | 24 (43.6) | 70 (23.3) | **0.003** |  | 17 (34.7) | 40 (14.0) | **0.001** |
| **Death (%)** | 37 (59.7) | 40 (12.2) | **0.001** |  | 36 (70.6) | 34 (11.5) | **0.001** |

AKI, acute kidney injury; MAP, mean arterial pressure; SOFA, Sequential Organ Failure
Assessment; CRP, C-reactive protein; LDH, lactate dehydrogenase; sCr, serum creatinine; CVD, cardiovascular disease; CKD, chronic kidney disease; BMI, body mass index; HFNO, high-flow nasal oxygen; NIV, noninvasive ventilation; ICU, intensive care unit.
§50% data missing

AKI patients were older (P≤0.001) than their non-AKI counterparts and they had a severe clinical course manifesting with a higher SOFA score (P≤0.001) and a more severe respiratory impairment (PO_2_/FO_2_, P≤0.001). This led to a longer hospital stay (P=0.001), complicated with a higher rate of ICU access and intubation for mechanical ventilation. As a result, a higher number of deaths (higher than 50%) occurred in the AKI subgroups: 59.7% during wave-1 and 70.6% during wave-2. At the end of the period of observation, 24% (wave-1) and 46.7% (wave-2) of survivors who experienced AKI were discharged with a not fully recovered kidney function (Table 1, supplementary material). The higher rate of unrecovered AKI during wave 2 was probably due to the shorter hospitalization compared to AKI wave-1 patients (P=0.008). Univariate and multivariate analysis of risk factors for AKI was performed in all AKI patients. Multivariate analysis showed that risk factors for AKI in COVID-19 disease were use of diuretics (HR=5.3; 95%CI:1.2-23.3; P=0.025) and cardiovascular disease (HR, 2.23; 95%CI:1.05-5.1; P=0.036) (Table1, Supplementary material).

## Discussion

The results of this study reported the trend of COVID-19-associated AKI in a high-risk area in Northern Italy. We found that the rate of new diagnosis of AKI in hospitalized patients was relatively steady (about 15%) across the “spring” and “fall” waves of COVID-19 that hit Europe in 2020. A similar incidence of AKI (15.8%) was surprisingly detected in summer, a period characterized by a low epidemiological risk and higher hospital bed capacity. These findings, albeit conceptually straightforward, provide new information on the epidemiology of AKI in COVID-19 patients.

AKI is a common complication of COVID-19 disease, which incidence is highly variable across countries. The causes of this heterogeneity are elusive, and no studies have evaluated until now the relationship of AKI with outbreaks. We cannot prove that the better healthcare organization of the second semester of 2020 has potentially reduced the episodes of AKI. Based on our data, the low rate and steady trend of AKI between waves could be interpreted epidemiologically as an indicator of high-quality nephrological care. From a practical point of view, the prevention measures of AKI were not different from those followed by other causes of AKI in critically ill patients[15,16]. These were based mainly on surveillance of kidney function, maintenance of normovolemia and avoidance of nephrotoxic agents.[17] We suppose that the close cooperation of nephrologists with other specialists contributed to maintaining a low rate AKI compared to other studies conducted in geographical zones with similar epidemiological risk.[18–20]

Overall, patients with AKI showed different demographic and clinical characteristics compared to the non-AKI patients. Kidney injury was indeed experienced by elder patients affected by a more severe disease than non-AKI patients. During the second wave, we noted a minor rate of ICU admissions and mechanical ventilation; conversely, this tendency was balanced by a compensative increase in non-invasive ventilation such as high-flow nasal oxygen (HFNO) and non-invasive ventilation (NIV), reflecting the prevalent opinions of that period of postponing invasive ventilation.**[21,22]**

According to the recent results showing a benefits of immunomodulators in COVID-19[23–26], a higher number of patients were treated with steroids during the wave-2, but the beneficial effects of steroids and anti-IL-6 in the prevention and mitigation of AKI remained unproved in our study. AKI was associated with classical risk factors such as diuretic and cardiovascular disease, suggesting dehydration or hemodynamic instability as a predisposing condition for kidney injury.

These data also underline the steady pathogenicity of the virus in inducing kidney injury in a setting not probably influenced by genetic variability of SARS-CoV-2[27]. Indeed, despite a steep learning curve on the management of the infected patients, morbidity and mortality of AKI was extremely higher than non-AKI patients, without any clear differences between the first and second wave. Further studies are required to investigate the consequences of new virulent SARS-CoV-2 variants and the effects of adjunctive therapies on the rate of AKI.

The potential underestimation of AKI events is the main limitation of this study. The lack of baseline creatinine before admission and daily urine output is unintended bias largely reported in other studies.

## Conclusion

In conclusion, our study shows a steady rate of AKI between the first two waves of COVID-19 (about 15%). AKI was associated with older age, higher level of inflammation, diuretics, and a consistent rate of non-recovery AKI.

## Data Availability

The data that support the findings of this study are available from the corresponding author upon reasonable reques

---

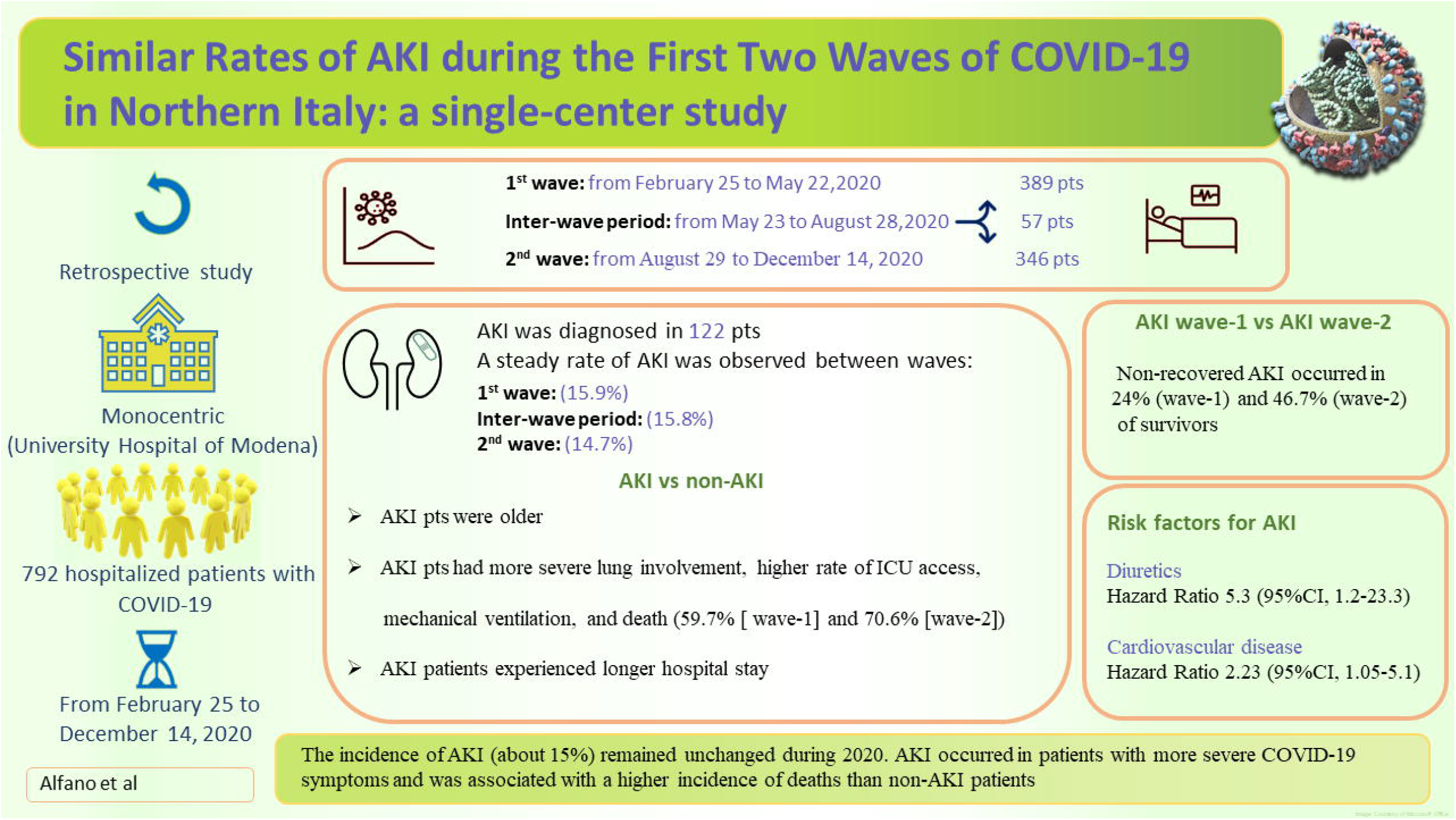

## Notes

**Conflicts of interest** All the authors have declared no competing interest

**Funding** This study was not funded

### Competing Interest Statement

The authors have declared no competing interest.

### Funding Statement

Not funded

### Author Declarations

The study was approved by the regional ethical committee of Emilia Romagna (n. 0013376/20)

### Summary of Updates

This version revised for figure 3.

## Bibliography

[1] COVID-19 situation update for the EU/EEA, as of 21 April 2021. Eur Cent Dis Prev Control n.d. https://www.ecdc.europa.eu/en/cases-2019-ncov-eueea (accessed April 21, 2021).

[2] Kellum JA, van Till JWO, Mulligan G. Targeting acute kidney injury in COVID-19. Nephrol Dial Transplant 2020;35:1652–62. https://doi.org/10.1093/ndt/gfaa231.

[3] Alessandri F, Pistolesi V, Manganelli C, Ruberto F, Ceccarelli G, Morabito S, et al. Acute Kidney Injury and COVID-19: A Picture from an Intensive Care Unit. Blood Purif 2021. https://doi.org/10.1159/000513153.

[4] Brienza N, Puntillo F, Romagnoli S, Tritapepe L. Acute Kidney Injury in Coronavirus Disease 2019 Infected Patients: A Meta-Analytic Study. Blood Purif 2021;50:35–41. https://doi.org/10.1159/000509274.

[5] Martínez-Rueda AJ, Álvarez RD, Méndez-Pérez RA, Fernández-Camargo DA, Gaytan-Arocha JE, Berman-Parks N, et al. Community-And Hospital-Acquired Acute Kidney Injury in COVID-19: Different Phenotypes and Dismal Prognosis. Blood Purif 2021. https://doi.org/10.1159/000513948.

[6] Fisher M, Neugarten J, Bellin E, Yunes M, Stahl L, Johns TS, et al. AKI in Hospitalized Patients with and without COVID-19: A Comparison Study. J Am Soc Nephrol 2020;31:2145–57. https://doi.org/10.1681/ASN.2020040509.

[7] Colaneri M, Sacchi P, Zuccaro V, Biscarini S, Sachs M, Roda S, et al. Clinical characteristics of coronavirus disease (COVID-19) early findings from a teaching hospital in Pavia, North Italy, 21 to 28 February 2020. Eurosurveillance 2020;25. https://doi.org/10.2807/1560-7917.ES.2020.25.16.2000460.

[8] Swiss Medical Weekly - Acute kidney injury in patients with COVID-19: a retrospective cohort study from Switzerland n.d. https://smw.ch/article/doi/smw.2021.20482 (accessed April 23, 2021).

[9] Portolés J, Marques M, López-Sánchez P, de Valdenebro M, Muñez E, Serrano ML, et al. Chronic kidney disease and acute kidney injury in the COVID-19 Spanish outbreak. Nephrol Dial Transplant 2020;35:1353–61. https://doi.org/10.1093/ndt/gfaa189.

[10] Vena A, Giacobbe DR, Di Biagio A, Mikulska M, Taramasso L, De Maria A, et al. Clinical characteristics, management and in-hospital mortality of patients with coronavirus disease 2019 in Genoa, Italy. Clin Microbiol Infect Off Publ Eur Soc Clin Microbiol Infect Dis 2020;26:1537–44. https://doi.org/10.1016/j.cmi.2020.07.049.

[11] Fu EL, Janse RJ, de Jong Y, van der Endt VHW, Milders J, van der Willik EM, et al. Acute kidney injury and kidney replacement therapy in COVID-19: a systematic review and meta-analysis. Clin Kidney J 2020;13:550–63. https://doi.org/10.1093/ckj/sfaa160.

[12] Palevsky PM. COVID-19 and AKI: Where Do We Stand? J Am Soc Nephrol 2021. https://doi.org/10.1681/ASN.2020121768.

[13] Kidney Disease: Improving Global Outcomes (KDIGO) Acute Kidney Injury Work Group. KDIGO Clinical Practice Guideline for Acute Kidney Injury. Kidney inter., Suppl. 2012; 2: 1–138. 2012:141.

[14] Cheng Y, Luo R, Wang X, Wang K, Zhang N, Zhang M, et al. The Incidence, Risk Factors, and Prognosis of Acute Kidney Injury in Adult Patients with Coronavirus Disease 2019. Clin J Am Soc Nephrol 2020;15:1394–402. https://doi.org/10.2215/CJN.04650420.

[15] Khwaja A. KDIGO Clinical Practice Guidelines for Acute Kidney Injury. Nephron Clin Pract 2012;120:c179–84. https://doi.org/10.1159/000339789.

[16] Rhodes A, Evans LE, Alhazzani W, Levy MM, Antonelli M, Ferrer R, et al. Surviving Sepsis Campaign: International Guidelines for Management of Sepsis and Septic Shock: 2016. Crit Care Med 2017;45:486–552. https://doi.org/10.1097/CCM.0000000000002255.

[17] Nadim MK, Forni LG, Mehta RL, Connor MJ, Liu KD, Ostermann M, et al. COVID-19-associated acute kidney injury: consensus report of the 25th Acute Disease Quality Initiative (ADQI) Workgroup. Nat Rev Nephrol 2020;16:747–64. https://doi.org/10.1038/s41581-020-00356-5.

[18] Hirsch JS, Ng JH, Ross DW, Sharma P, Shah HH, Barnett RL, et al. Acute kidney injury in patients hospitalized with COVID-19. Kidney Int 2020;98:209–18. https://doi.org/10.1016/j.kint.2020.05.006.

[19] Fisher M, Neugarten J, Bellin E, Yunes M, Stahl L, Johns TS, et al. AKI in Hospitalized Patients with and without COVID-19: A Comparison Study. J Am Soc Nephrol JASN 2020;31:2145–57. https://doi.org/10.1681/ASN.2020040509.

[20] Wan YI, Bien Z, Apea VJ, Orkin CM, Dhairyawan R, Kirwan CJ, et al. Acute Kidney Injury in COVID-19: multicentre prospective analysis of registry data. Clin Kidney J 2021. https://doi.org/10.1093/ckj/sfab071.

[21] Bonnet N, Martin O, Boubaya M, Levy V, Ebstein N, Karoubi P, et al. High flow nasal oxygen therapy to avoid invasive mechanical ventilation in SARS-CoV-2 pneumonia: a retrospective study. Ann Intensive Care 2021;11:37. https://doi.org/10.1186/s13613-021-00825-5.

[22] Tobin MJ, Laghi F, Jubran A. Caution about early intubation and mechanical ventilation in COVID-19. Ann Intensive Care 2020;10:78. https://doi.org/10.1186/s13613-020-00692-6.

[23] Salton F, Confalonieri P, Meduri GU, Santus P, Harari S, Scala R, et al. Prolonged Low-Dose Methylprednisolone in Patients With Severe COVID-19 Pneumonia. Open Forum Infect Dis 2020;7:ofaa421. https://doi.org/10.1093/ofid/ofaa421.

[24] WHO Rapid Evidence Appraisal for COVID-19 Therapies (REACT) Working Group, Sterne JAC, Murthy S, Diaz JV, Slutsky AS, Villar J, et al. Association Between Administration of Systemic Corticosteroids and Mortality Among Critically Ill Patients With COVID-19: A Meta-analysis. JAMA 2020;324:1330–41. https://doi.org/10.1001/jama.2020.17023.

[25] RECOVERY Collaborative Group, Horby P, Lim WS, Emberson JR, Mafham M, Bell JL, et al. Dexamethasone in Hospitalized Patients with Covid-19. N Engl J Med 2021;384:693– 704. https://doi.org/10.1056/NEJMoa2021436.

[26] Guaraldi G, Meschiari M, Cozzi-Lepri A, Milic J, Tonelli R, Menozzi M, et al. Tocilizumab in patients with severe COVID-19: a retrospective cohort study. Lancet Rheumatol 2020;0. https://doi.org/10.1016/S2665-9913(20)30173-9.

[27] Lo Menzo S, Marinello S, Biasin M, Terregino C, Franchin E, Crisanti A, et al. The first familial cluster of the B.1.1.7 variant of SARS-CoV-2 in the northeast of Italy. Infection 2021:1–5. https://doi.org/10.1007/s15010-021-01609-6.

